# High visit-to-visit cholesterol variability predicts heart failure and adverse cardiovascular events: a population-based cohort study

**DOI:** 10.1101/2021.12.10.21267624

**Authors:** Jeffrey Shi Kai Chan, Danish Iltaf Satti, Yan Hiu Athena Lee, Jeremy Man Ho Hui, Teddy Tai Loy Lee, Oscar Hou In Chou, Abraham Ka Chung Wai, Ana Ciobanu, Ying Liu, Tong Liu, Qingpeng Zhang, Bernard Man Yung Cheung, Jiandong Zhou, Gary Tse

## Abstract

**Background:** Dyslipidaemia is associated with adverse cardiovascular outcomes. However, the long-term prognostic value of visit-to-visit cholesterol variability for the risks of heart failure (HF) is uncertain. We investigated the associations between cholesterol variability and the risk of HF and adverse cardiovascular events.

**Methods:** This retrospective cohort study included patients attending family medicine clinics in Hong Kong during 2000-2003 with follow-up until 2019. Patients with at least three sets of blood cholesterol (low-density (LDL-C) and high-density (HDL-C) lipoprotein cholesterol) levels available at different visits were included. Patients with prior HF, myocardial infarction (MI), use of HF medications, and pregnancy were excluded. Visit-to-visit variability was calculated using standard deviation and coefficient of variation (CV). The primary outcome was HF. The secondary outcomes were cardiovascular mortality, and myocardial infarction.

**Results:** A total of 5662 patients were included (2152 males; mean age 63.3±12.4 years; mean follow-up 15.3±4.6 years). Higher variability of HDL-C (hazard ratio (HR) for CV: 13.757 [6.261, 30.226], p<0.0001) predicted new-onset HF. Higher variability of LDL-C (HR for CV: 3.885 [1.942, 7.775], p=0.0001) and HDL-C (HR for CV: 39.118 [13.583, 112.657], p<0.0001) predicted higher risk of MI, but not cardiovascular mortality. These associations remained significant in patients without baseline usage of lipid-lowering medication(s) (N=4068), but were all insignificant in patients with baseline usage of lipid-lowering medication(s) (N=1594).

**Conclusion:** Higher visit-to-visit cholesterol variability was varyingly associated with significantly increased long-term risks of HF and adverse cardiovascular events. Such associations may be negated by using lipid-lowering medication(s).

## Introduction

Dyslipidaemia has been associated with elevated risks of cardiovascular diseases, with high LDL-C and low HDL-C levels generally regarded to be detrimental.(1,2) Among cardiovascular diseases, heart failure is one of the most important public health problems associated with significant mortality, morbidity, and healthcare expenditures. It had been declared as an emerging pandemic in 1997,(3) and with global cases rising from 33.5 million in 1990 to 64.3 million in 2017, strategies are needed to prevent underlying causes to reduce future burden of this disease.(4) Nonetheless, despite the well-established roles of cholesterol in atherosclerosis and thus ischaemic heart disease, less is known about the relationship between cholesterol levels and heart failure.(5) Studies in this area have had contradicting results, with some studies finding significant relationships, and others finding none, particularly in the general population.(6,7) Additionally, most studies have focused on the LDL-C and HDL-C levels as mean levels or point estimates, despite lipid levels being known display significant intra-personal variations.(8)

Meanwhile, studies have suggested that visit-to-visit lipid variability might have incremental prognostic values for adverse cardiovascular and cerebrovascular events, in addition to mean cholesterol levels.(9–11) A post-hoc analysis of the TNT trial showed that in selected patients with coronary artery disease, every 1-SD increase in LDL-C variability was associated with 10-20% increase in the risks of major adverse cardiovascular events, independent of adherence to trial medications.(12) Meanwhile, Han *et al*. showed that, in a Korean cohort taken from the general population, coexistence of lower mean and higher variability of HDL-C was associated with 23-47% increases in the risks of various adverse cardiovascular events.(13) Despite a growing body of evidence suggesting that cholesterol variability may predict the risks of various cardiovascular conditions, evidence pertaining to heart failure remains scarce and, where existent, only explored the prognostic implications of the variability of total cholesterol and not that of LDL-C and HDL-C separately.(13) As such, the present study aimed to investigate the association between LDL-C and HDL-C variabilities and the risk of heart failure and adverse cardiovascular outcomes.

## Methods

This retrospective cohort study was approved by The Joint Chinese University of Hong Kong – New Territories East Cluster Clinical Research Ethics Committee and Institutional Review Board of the University of Hong Kong/Hospital Authority Hong Kong West Cluster. It was performed in accordance with the Declaration of Helsinki. Requirement for individual patient consent was waived due to the use of retrospective deidentified data. All data underlying the results of this study are available upon reasonable request to the corresponding authors.

### Source of data and study population

All data were retrieved from the Clinical Data Analysis and Reporting System (CDARS), a territory-wide database which records key demographic, diagnostic, procedural and medication records of all patients attending any public healthcare institution in Hong Kong. All diagnoses are coded in the *International Classification of Diseases Ninth revision* (ICD-9) or *Tenth revision* (ICD-10) codes depending on the year of entry, with the codes entered by treating clinicians. Owing to the follow-up period of the current study population, all diagnoses hereby mentioned were identified by ICD-9 codes. The system was linked to the Hong Kong Death registry, a population-based governmental registry of the death records of all Hong Kong citizens, from which mortality data may be obtained. CDARS and the associated mortality data have previously been used for research both by our team and other teams in Hong Kong.(14–16)

The inclusion criteria were adult patients (at least 18 years old) who attended a family medicine clinic in Hong Kong between 1^st^ January 2000 to 31^st^ December 2003; only those who had at least three sets of fasting lipid profiles, which at least included LDL-C and HDL-C, were included. The exclusion criteria were prior diagnosis of heart failure, prior myocardial infarction, using heart failure medications at baseline, pregnancy at baseline, known infection by human immunodeficiency virus, and patients who had less than three sets of lipid profiles available.

### Follow-up, outcomes, and study variables

All patients were followed up until 31^st^ December 2019. The primary outcome was heart failure. The secondary outcomes were cardiovascular mortality, and myocardial infarction. All outcomes were identified using ICD-9 codes.

Key demographic measures of all included patients were obtained, including age at baseline, and biological sex. Other comorbidities were also recorded, including hypertension, diabetes mellitus, atrial fibrillation, chronic obstructive pulmonary disease, ischaemic heart disease, peripheral vascular disease, stroke or transient ischaemic attack, and dementia or Alzheimer’s disease. The ICD-9 codes used for identifying all relevant diagnoses in this study are listed in **Supplementary Table 1**. The usage of any lipid-lowering medications was recorded as well.

The fasting levels of LDL-C and HDL-C from all available test records during the study period were recorded, from which the mean, standard deviation (SD), and coefficient of variation (CV) which was defined as SD divided by mean, were calculated. Both SD and CV were used as markers of variability.

### Statistical analyses

Continuous variables were expressed as mean±SD. Student’s t test was used to compare continuous variables between subgroups, while Fisher’s exact test was used to compare binary variables. Cox proportional hazards regression analysis was used to assess the associations between the mean and variability of LDL-C and HDL-C and the outcomes.

Univariable Cox regression was performed for all the above demographic and comorbidity variables to identify covariates that had substantial effects on the outcomes, as defined by p<0.010. All such variables were subsequent used in multivariable Cox regression analyses to adjust the associations between the above cholesterol measures and the outcomes. In addition to performing Cox regression analyses on the overall study cohort, an *a priori* subgroup analysis by the usage of any lipid-lowering medication was performed with the same analytical methods. Hazard ratios (HR) with 95% confidence intervals (CI) were used as the summary statistics for the results from Cox regression analyses. The marginal effects of lipid profiles and their variability measures were also calculated. The partial derivatives with respect to lipid profile and their variability measures in the Cox regression model for each unit in the data were identified, which provide better understanding about the unit-specific partial derivatives of them over the cohort. The marginal effects were visualized with 95% confidence intervals to facilitate interpretation of their associations with adverse study outcomes.

All p values were two-sided, and p<0.05 were considered statistically significant. All statistical analyses and visualizations were performed on SPSS version 25.0 (IBM Corp, New York, USA), Stata (Version 13.0) RStudio software (Version: 1.1.456) and Python (Version: 3.6).

## Results

The flow chart of this study is shown in **Figure 1**. A total of 155,065 patients were identified by the inclusion criteria. After excluding 1062 patients with prior diagnosis of heart failure, 192 with prior myocardial infarction, 3866 with baseline usage of heart failure medications, 152 with pregnancy, 11 with known infection by human immunodeficiency virus, and 144,120 patients without at least three sets of lipid profile test results, a total of 5662 patients were included in this study (2152 males; mean age 63.3±12.4 years) with a mean of 13±9 sets of lipid profile results available for analysis. Of the included patients, 1594 patients (28.2%) were taking lipid-lowering medications at baseline. The baseline characteristics of the included patients were summarized in **Table 1**. Over a mean follow-up period of 15.3±4.6 years, the primary outcome of heart failure occurred in 1196 (21.1%) patients, while the secondary outcomes of cardiovascular mortality occurred in 548 (9.68%) and 656 (11.6%) patients respectively.

**Table 1.**
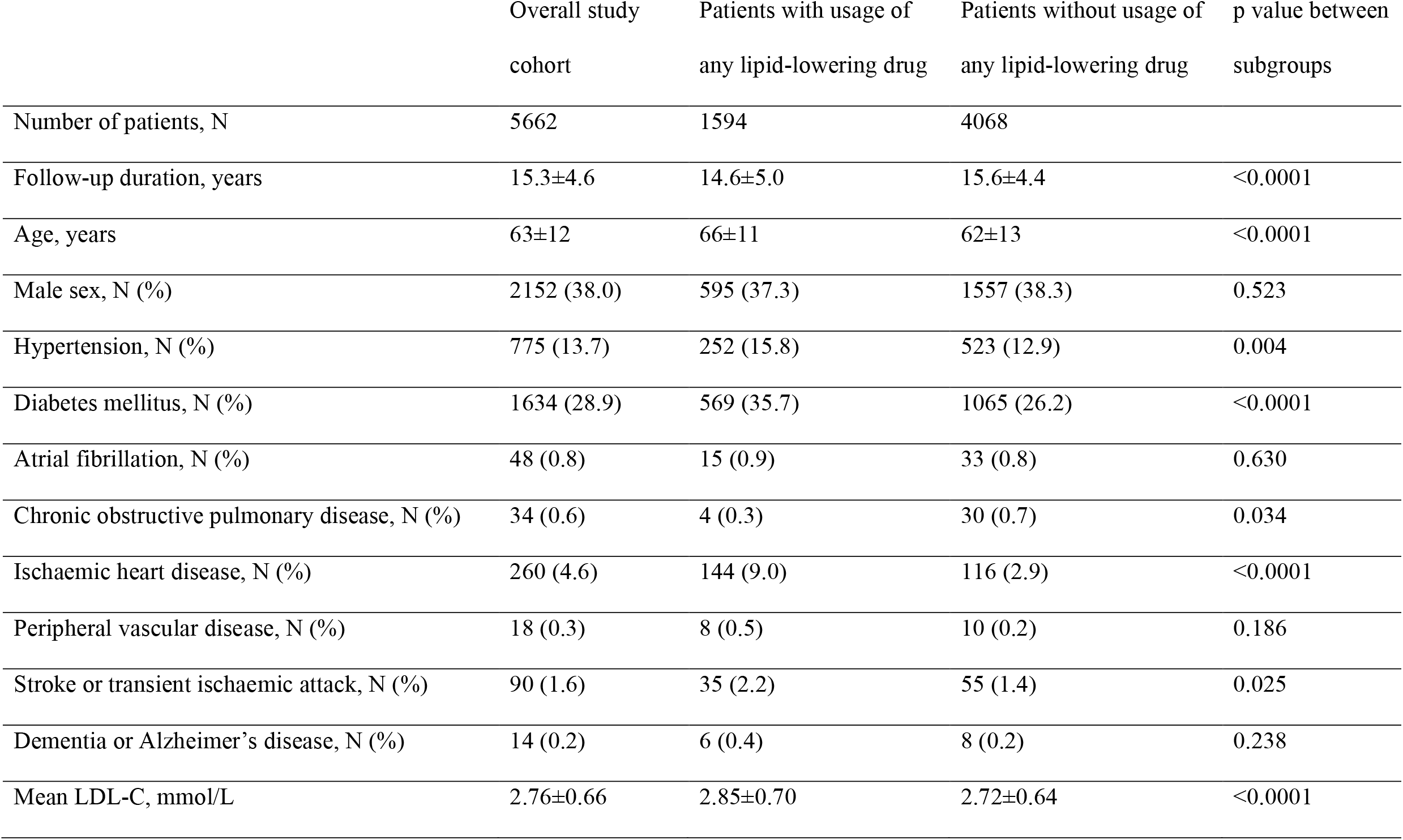
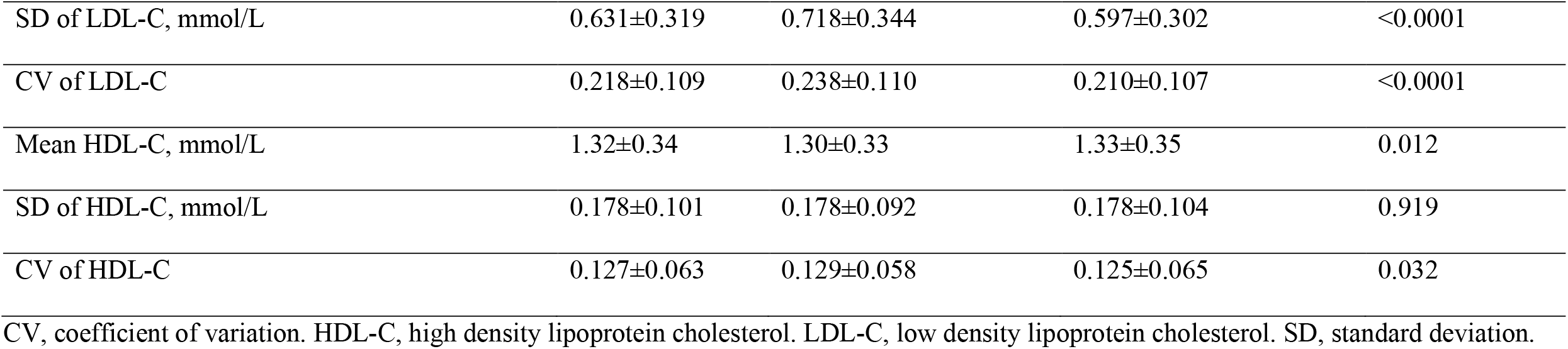
Baseline characteristics of included patients.

**Table 2.** Results of multivariable Cox regression analysis for the overall study cohort.

|  | Hazard ratio [95% confidence interval], p value |  |  |
| --- | --- | --- | --- |
|  | New-onset heart failure <sup>a</sup> | Cardiovascular mortality <sup>b</sup> | Myocardial infarction <sup>c</sup> |
| Mean LDL-C | 0.980 [0.896, 1.073], p=0.668 | 1.396 [1.229, 1.587], p<0.0001 | 0.956 [0.843, 1.084], p=0.483 |
| SD of LDL-C | 1.194 [1.001, 1.425], p=0.049 | 1.025 [0.785, 1.338], p=0.858 | 1.503 [1.187, 1.902], p=0.001 |
| CV of LDL-C | 1.465 [0.868, 2.475], p=0.153 | 0.231 [0.102, 0.521], p<0.0001 | 3.885 [1.942, 7.775], p=0.0001 |
| Mean HDL-C | 0.563 [0.470, 0.676], p<0.0001 | 0.533 [0.404, 0.703], p<0.0001 | 0.460 [0.355, 0.596], p<0.0001 |
| SD of HDL-C | 1.966 [1.193, 3.238], p=0.008 | 1.033 [0.458, 2.327], p=0.938 | 2.777 [1.405, 5.489], p=0.003 |
| CV of HDL-C | 13.757 [6.261, 30.226], p<0.0001 | 1.662 [0.461, 5.996], p=0.438 | 39.118 [13.583, 112.657], p<0.0001 |

CV, coefficient of variation. HDL-C, high density lipoprotein cholesterol. LDL-C, low density lipoprotein cholesterol. SD, standard deviation.
<sup>a</sup> Adjusted for age, hypertension, diabetes mellitus, atrial fibrillation, chronic obstructive pulmonary disease, ischaemic heart disease, peripheral vascular disease, stroke or transient ischaemic attack, and dementia or Alzheimer's disease
<sup>b</sup> Adjusted for age, sex, hypertension, diabetes mellitus, atrial fibrillation, ischaemic heart disease, peripheral vascular disease, stroke or transient ischaemic attack, and dementia or Alzheimer's disease
<sup>c</sup> Adjusted for age, sex, hypertension, diabetes mellitus, chronic obstructive pulmonary disease, ischaemic heart disease, peripheral vascular disease, and stroke or transient ischaemic attack

**Figure 1.**
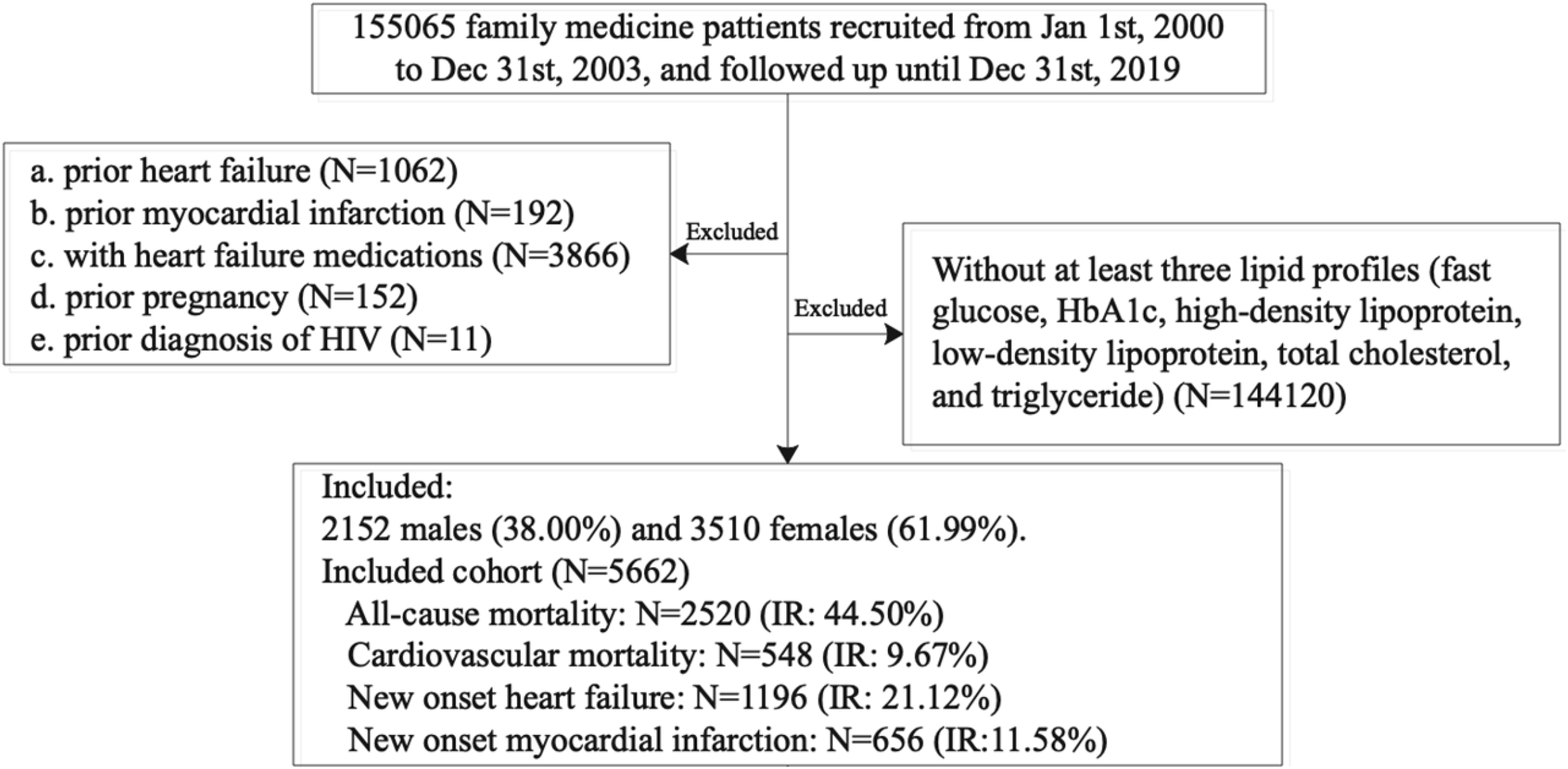
Study flow chart.

### Associations between cholesterol measures and outcomes

Univariable and multivariable Cox regression was performed, with their respective results shown in **Supplementary Table 2** and **Table 2**, respectively. Higher HDL-C variability, as measured by SD and CV, was associated with significantly higher risk of heart failure (adjusted HR 1.966 [1.193, 3.238], p=0.008, and 13.757 [6.261, 30.226], p<0.0001, respectively). Although higher SD of LDL-C was associated with higher risk of heart failure (adjusted HR 1.194 [1.001, 1.425], p=0.049), no significant association was observed between CV of LDL-C and risk of heart failure (p=0.153). Lower mean HDL-C was associated with significantly higher risk of heart failure (adjusted HR 0.563 [0.470, 0.676], p<0.0001), while mean LDL-C was not significantly associated with the risk of heart failure. These relationships are also shown in **Figure 2**, which demonstrates strong relationships between HDL-C measures and risk of heart failure, but none for LDL-C measures.

**Figure 2.**
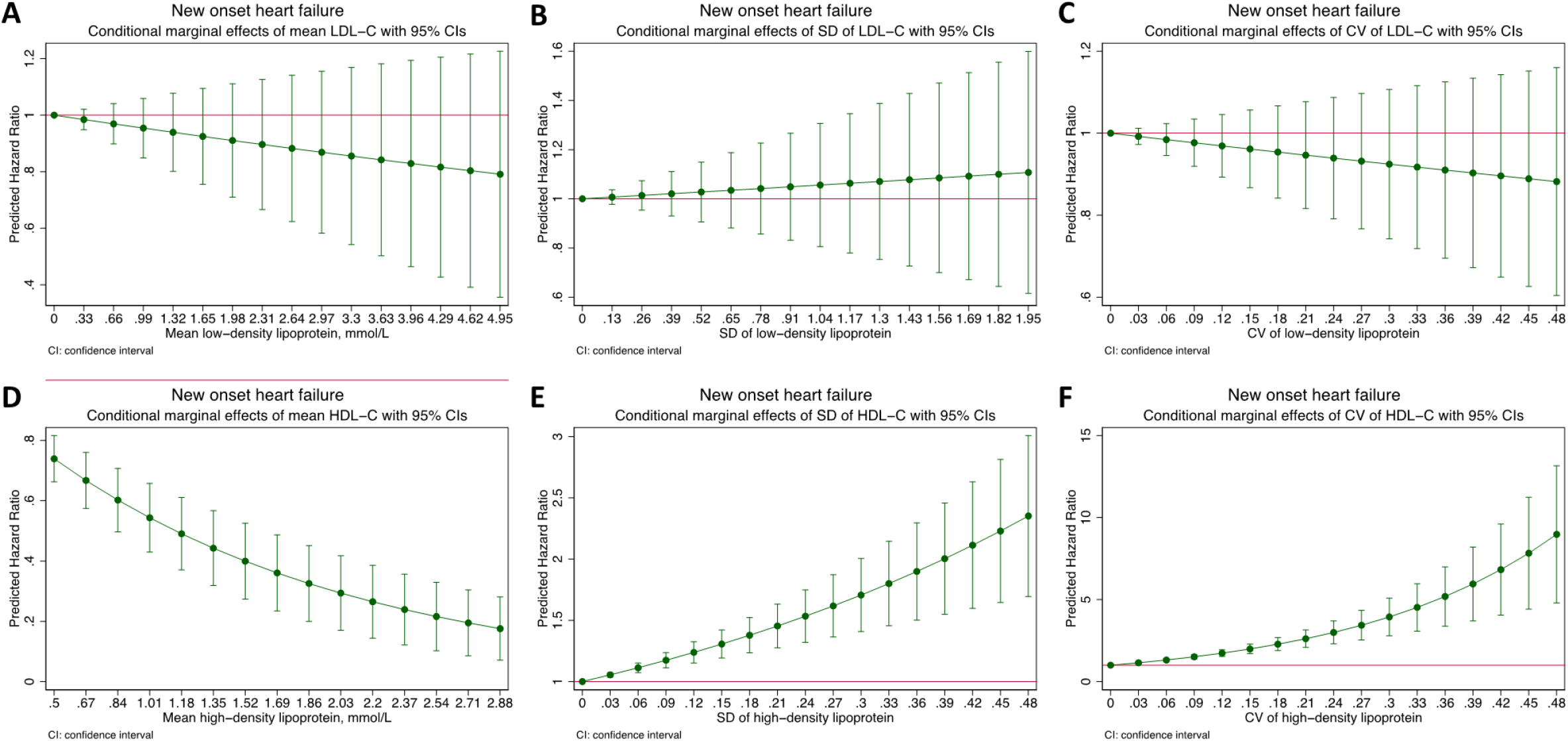
Marginal effects of the mean low density lipoprotein cholesterol (LDL-C; **A**), standard deviation (SD) of LDL-C (**B**), coefficient of variation (CV) of LDL-C (**C**), mean high density lipoprotein cholesterol (HDL-C; **D**), SD of HDL-C (**E**), and CV of HDL-C (**F**) on the risk of new-onset heart failure.

Meanwhile, there were no significant associations between the SD of LDL-C (p=0.858) or either measure of HDL-C variability (p=0.938 for SD and p=0.438 for CV) and cardiovascular mortality. Higher mean LDL-C (adjusted HR 1.396 [1.229, 1.587], p<0.0001) and lower mean HDL-C (adjusted HR 0.533 [0.404, 0.703], p<0.0001) were both associated with higher risk of cardiovascular mortality; the inverse correlation between the CV of LDL-C and cardiovascular mortality (p<0.0001) was thus likely driven by the positive correlation between mean LDL-C and cardiovascular mortality. These observations are also shown graphically in **Figure 3**.

**Figure 3.**
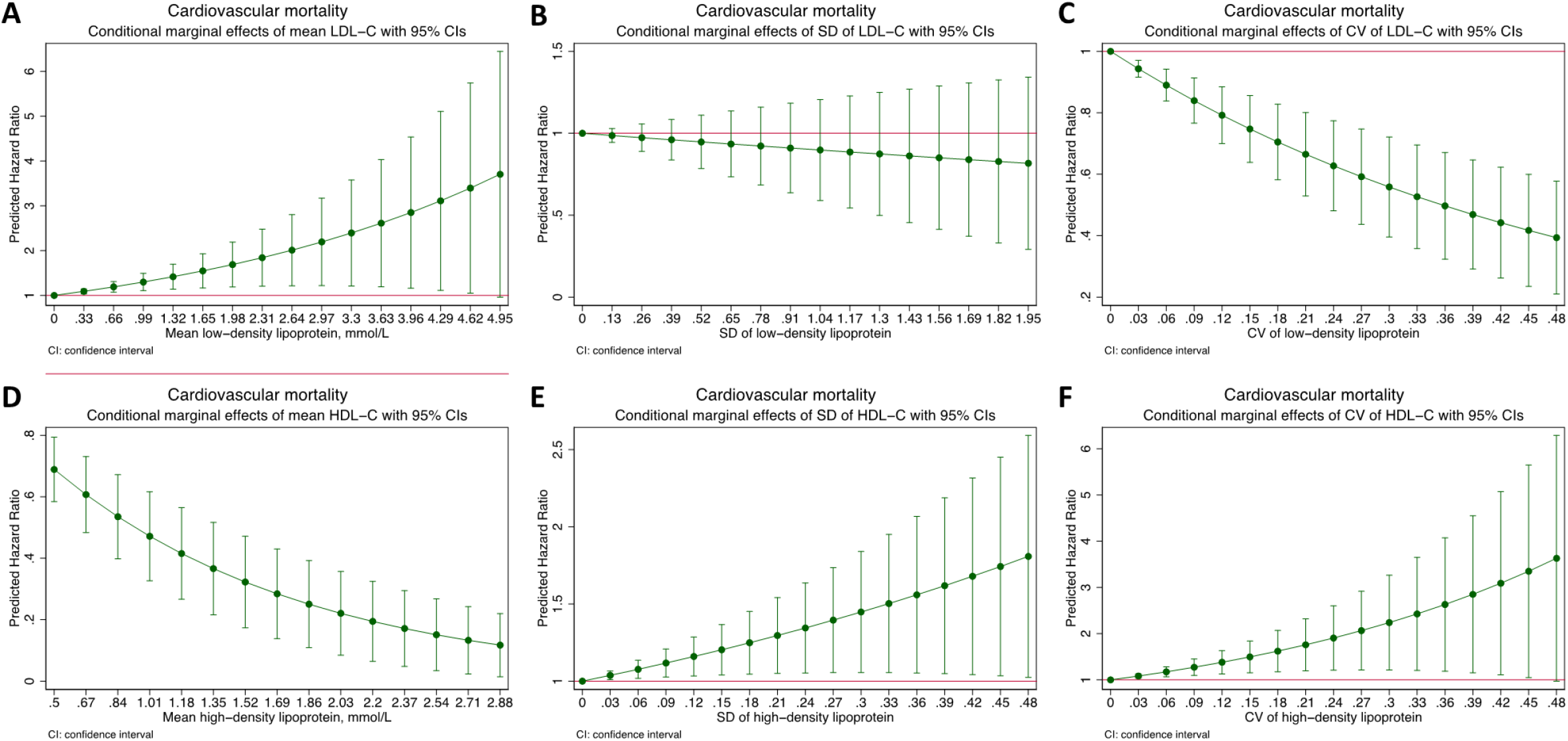
Marginal effects of the mean low density lipoprotein cholesterol (LDL-C; **A**), standard deviation (SD) of LDL-C (**B**), coefficient of variation (CV) of LDL-C (**C**), mean high density lipoprotein cholesterol (HDL-C; **D**), SD of HDL-C (**E**), and CV of HDL-C (**F**) on the risk of cardiovascular mortality.

Furthermore, higher variability of both LDL-C (adjusted HR 1.503 [1.187, 1.902], p=0.001 for SD, and 3.885 [1.942, 7.775], p=0.0001 for CV) and HDL-C (adjusted HR 2.777 [1.405, 5.489], p=0.003 for SD, and 39.118 [13.583, 112.657], p<0.0001 for CV) was associated with higher risk of myocardial infarction. While lower mean HDL-C was associated with higher risk of myocardial infarction (adjusted HR 0.460 [0.355, 0.596], p<0.0001), mean LDL-C was not observed to be significantly associated with the risk of myocardial infarction. These observations are also shown graphically in **Figure 4**

**Figure 4.**
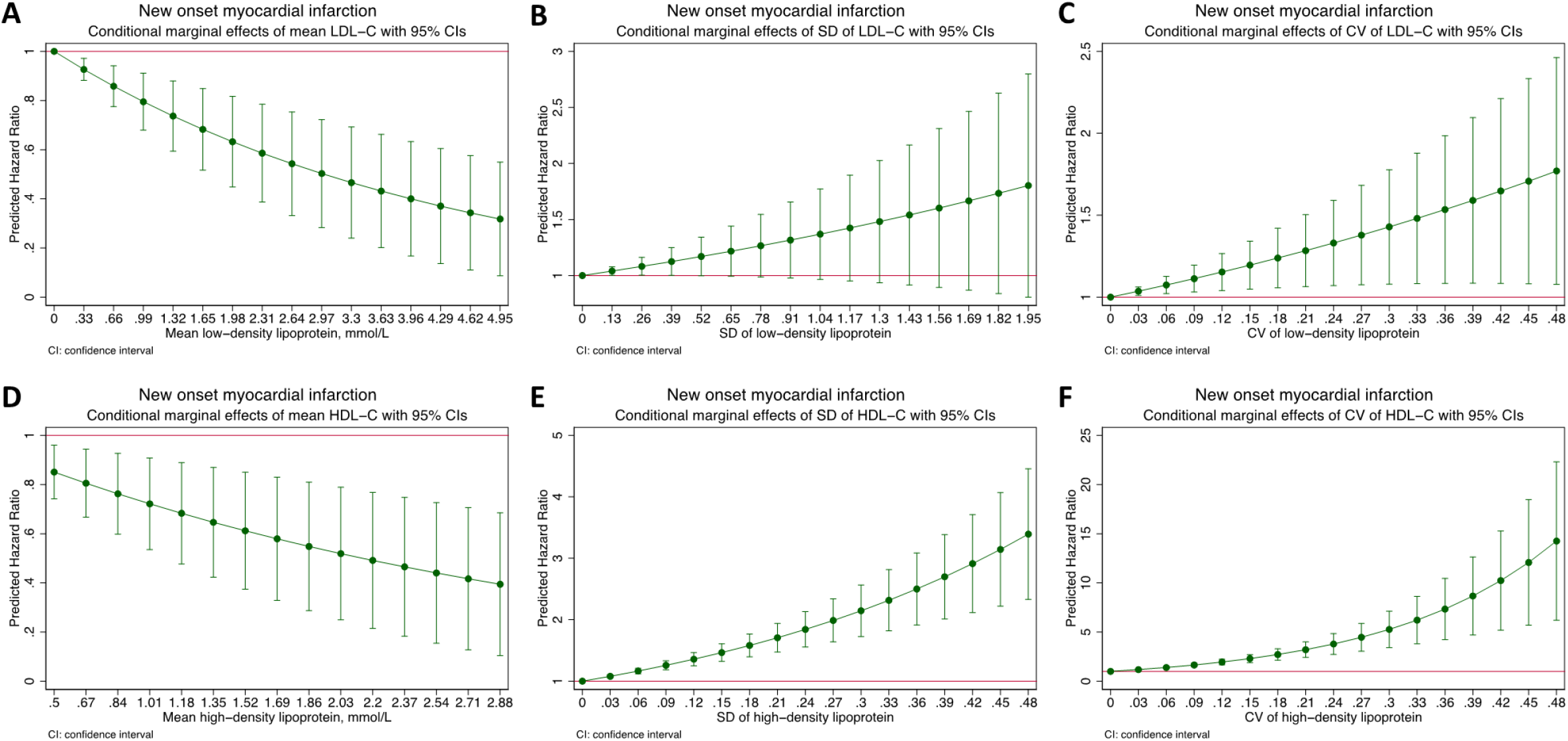
Marginal effects of the mean low density lipoprotein cholesterol (LDL-C; **A**), standard deviation (SD) of LDL-C (**B**), coefficient of variation (CV) of LDL-C (**C**), mean high density lipoprotein cholesterol (HDL-C; **D**), SD of HDL-C (**E**), and CV of HDL-C (**F**) on the risk of new-onset myocardial infarction.

### Subgroup analysis: patients without baseline usage of lipid-lowering medication(s)

The baseline characteristics of both subgroups are summarized in **Table 1**. The results of univariable Cox regression for both subgroups are summarized in **Supplementary Table 3 and 4**, while the corresponding multivariable results are summarized in **Table 3**. Patients without baseline usage of lipid-lowering medication(s) were slightly younger (62±13 years versus 66±11 years, p<0.0001) and had slightly mean longer follow-up duration (15.6±4.4 years versus 14.6±5.0 years). Additionally, among these patients, lower prevalence of diabetes mellitus (p<0.0001) and ischaemic heart disease (p<0.0001) were observed. Lower mean, SD, and CV of LDL-C (p<0.0001 for all), and higher mean (p=0.012) and lower CV (p=0.032) of HDL-C were also observed.

**Table 3.**
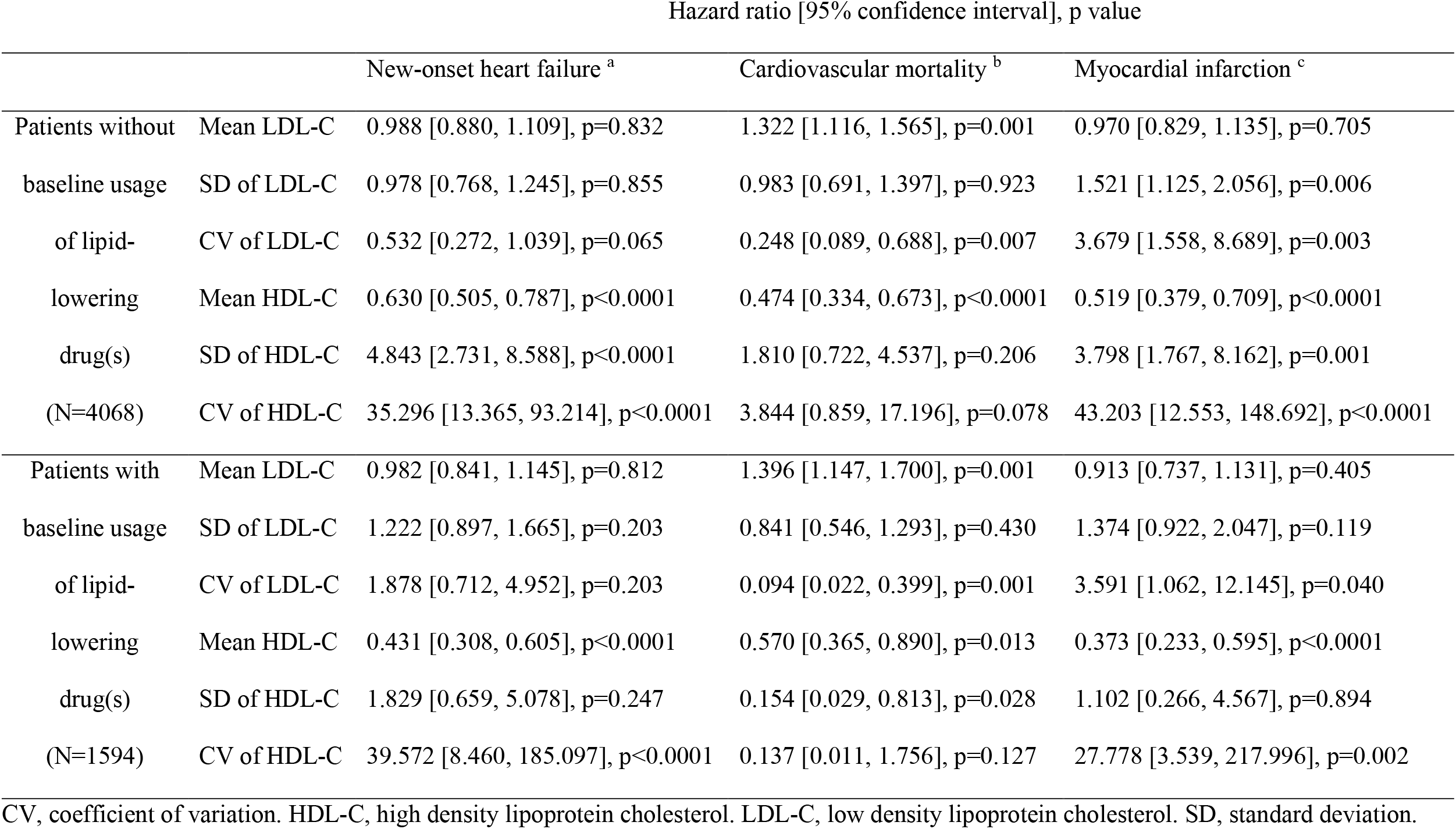
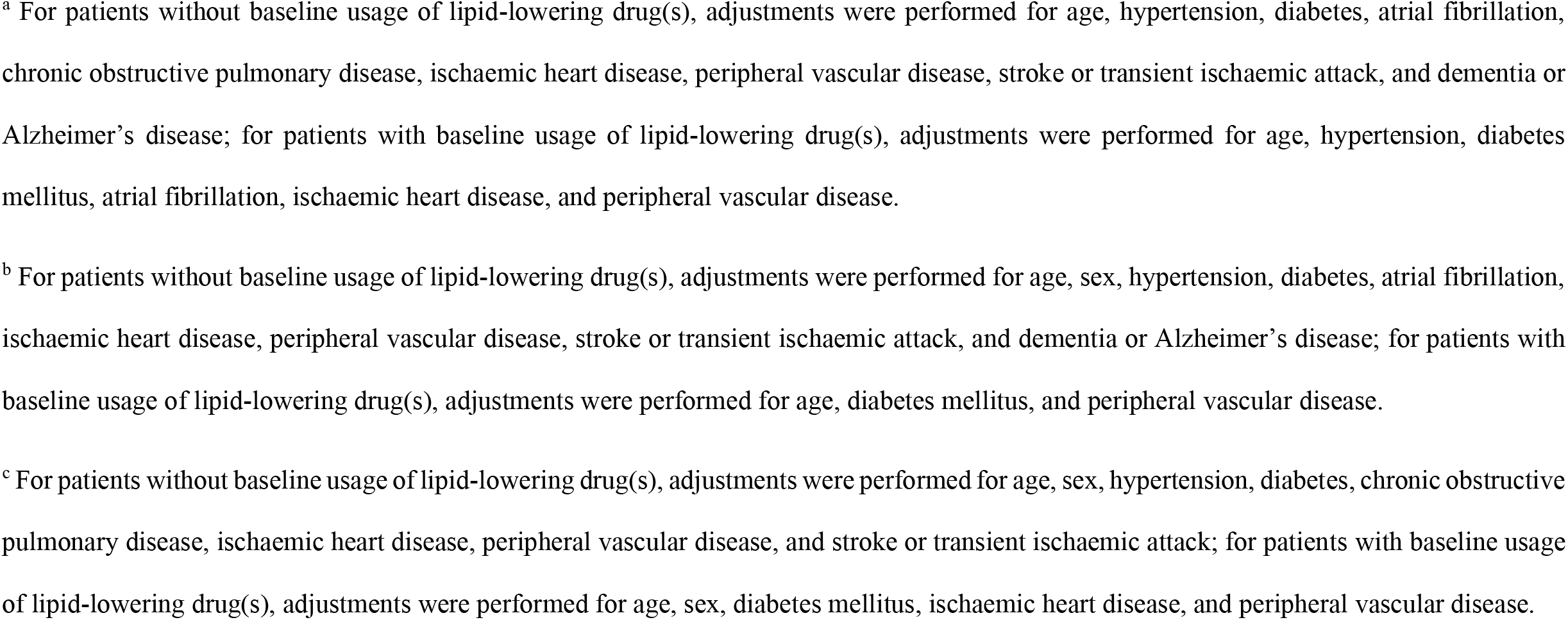
Results of multivariable Cox regression analysis for subgroups of patients stratified by baseline usage of lipid-lowering drug(s).

In patients who were not using lipid-lowering medication(s) at baseline (N=4068), the observed associations between LDL-C and HDL-C measures and outcomes were similar to those made in the overall study cohort: lower mean HDL-C and higher HDL-C variability were associated with higher risk of heart failure, higher mean LDL-C and lower mean HDL-C were associated with higher risk of cardiovascular mortality, and higher LDL-C or HDL-C variability and lower mean HDL-C were associated with higher risk of myocardial infarction.

### Subgroup analysis: patients with baseline usage of lipid-lowering medication(s)

In contrast, in patients who were using lipid-lowering medication(s) at baseline (N=1594), LDL-C and HDL-C variability were generally not observed to be significantly prognostic. While lower mean HDL-C was associated with higher risk of heart failure (adjusted HR 0.431 [0.308, 0.605], p<0.0001), no association was observed between SD of HDL-C and the risk of heart failure (adjusted HR 1.829 [0.659, 5.078], p=0.247), which suggested that the positive correlation between CV of HDL-C and heart failure was driven by the above correlation between mean HDL-C and heart failure risk. There were no significant associations between any LDL-C measures and the risk of heart failure.

Higher mean LDL-C (adjusted HR 1.396 [1.147, 1.700], p=0.001) and lower mean HDL-C (adjusted HR 0.570 [0.365, 0.890], p=0.013) were associated with higher risk of cardiovascular mortality, consistent with the observations made in the overall cohort analysis. The lack of association between the SD of LDL-C and risk of cardiovascular mortality suggested the inverse correlation between the CV of LDL-C and the said risk was driven by the abovementioned positive correlation for mean LDL-C. By contrast, the SD of HDL-C was inversely correlated to the risk of cardiovascular mortality, while its CV displayed no significant associations with the said risk. The inverse correlation for the SD of HDL-C was thus likely driven by the aforementioned inverse correlation for mean HDL-C, since lower HDL-C levels would proportionally lead to lower values in SD but not CV.

Lastly, there was no significant association between the SD of HDL-C and the risk of myocardial infarction. Lower mean HDL-C (adjusted HR 0.373 [0.233, 0.595], p<0.0001), but not mean LDL-C, was associated with higher risk of myocardial infarction, consistent with the observations in the overall study cohort analysis. These suggested that the positive correlation between the CV of HDL-C and risk of myocardial infarction in these patients was again due to the inverse correlation between mean HDL-C and myocardial infarction risk.

## Discussion

In this population-based cohort study, we showed that visit-to-visit LDL-C and HDL-C variability had important but varying associations with the long-term risks of heart failure and adverse cardiovascular outcomes. Such associations appeared to be negated by the use of lipid-lowering drugs. To the best of our knowledge, this is the first study to demonstrate a link between cholesterol variability and the risk of heart failure.

### Potential underlying mechanisms

While the exact mechanisms underlying the observed associations between cholesterol variability and cardiovascular risks remain unclear, several potential explanations exist. One such possibility pertains to the effects of lipid levels on atherosclerotic plaques. Constant fluctuations in serum lipid levels may alter the structure and composition of atherosclerotic plaques, inducing recurrent dissolution and crystallization of cholesterol.(17) In turn, these may lead to plaque volume expansion and plaque rupture, which are major mechanisms underlying myocardial infarction.(17) This has been confirmed by Nakano *et al* using optical coherence tomography in patients with acute coronary syndrome.(18) Progression of subclinical atheroma may also be accelerated by higher LDL-C variability, with LDL-C variability having been observed to directly correlate to carotid media intima thickness in patients with type 2 diabetes mellitus, as well as correlating to coronary atheroma progression in patients with coronary artery disease.(19,20) Additionally, higher LDL-C variability may lead to new plaque formation and atherosclerosis via endothelial dysfunction,(21,22) as well as causing plaque instability by impairing cholesterol efflux from peripheral tissues and macrophages.(23) These effects on atherosclerotic plaques may have contributed significantly to our findings, particularly the observation that higher variabilities of LDL-C and HDL-C both predicted the risk of myocardial infarction. These may also explain the variability measures’ lack of prognostic value in patients who were taking lipid-lowering medication(s), since some lipid-lowering drugs, such as statins, possess plaque-stabilizing properties.(24) Such properties may have negated the detrimental effects of cholesterol variability on atherosclerotic plaques, thereby leading to our observed lack of prognostic power.

Additionally, some exploratory studies have suggested a possible link between cholesterol variability and inflammation, with HDL-C variability demonstrating particularly strong associations.(25,26) Given that inflammation plays important roles in the pathogenesis of heart failure,(27,28) this is a possible mechanism mediating the observed association between elevated HDL-C variability and elevated risk of heart failure. This was further supported by a recent study by Park *et al* who observed that both elevated C-reactive protein levels and HDL-C variability were associated with the risk of subclinical left ventricular diastolic dysfunction.(29) The anti-inflammatory effects of some lipid-lowering medications, such as statins and fibrates,(30–32) would then explain the lack of association between HDL-C variability and risk of heart failure in patients who were using lipid-lowering medication(s). It is important to note that in Hong Kong, the most common cause of heart failure is hypertensive cardiomyopathy, instead of ischaemic cardiomyopathy which is the most common cause of heart failure worldwide.(33) This implies that in the present cohort, the aforementioned changes in atherosclerotic progression and plaque behaviour may not be adequate to explain the association between HDL-C variability and risk of heart failure; this was also suggested by the unique association for HDL-C variability, but not LDL-C variability which have been demonstrated previously to have strong effects on atherosclerotic plaques. Additionally, hypertensive cardiomyopathy most commonly results in diastolic dysfunction and heart failure with preserved ejection fraction. Therefore, the unusual epidemiology of heart failure in Hong Kong implied that the aforementioned findings by Park *et al* may be especially relevant in the present study.(29) Overall, inflammation may be reasonably postulated to be a mediating factor underlying our findings, and further studies are required to delineate the relationship between cholesterol variability, inflammation, and heart failure more definitively.

### What we knew and what is new

Previous studies have demonstrated some relationships between cholesterol variability and the risk of selected adverse cardiovascular outcomes. Looking at both specific patient populations(34) and the general population,(35) the outcomes of interest have been limited to mortality, myocardial infarction, and stroke, the latter two of which were both predominantly atherosclerotic events. To the best of our knowledge, the present study is the first investigation focusing on the relationships between cholesterol variability and the risk of heart failure. With our results in agreement with the existing literature, our findings build on and expand the existing understanding of the prognostic value of cholesterol variability. The pathogenesis of heart failure involves complex mechanisms beyond atherosclerosis,(36) and, as aforementioned, our observation of significant associations between HDL-C variability and the risk of heart failure may signal new opportunities of understanding the links between HDL-C variability and other pathogenic mechanisms such as inflammation, as well as the pathophysiology of heart failure.

In addition, most previous studies were limited to less than 10 years of median or mean follow-up duration.(34,35,37,38) In contrast, making use of a population-based database, the present cohort was followed up for a mean duration of more than 15 years. Our findings thus demonstrated the long-term prognostic value of cholesterol variability, which, when considered together with the existing literature, implies that cholesterol variability has important prognostic implications regardless of timeframe.

### Clinical implications and the future

This study provides strong evidence supporting the inclusion of cholesterol variability as part of routine cardiovascular risk stratification assessments. First, since lipid levels are known to fluctuate considerably,(8) we hope that our findings raise clinicians’ awareness of the importance of cholesterol variability, and reduce the likelihood of clinically important variability being dismissed as mere measurement errors. Second, since we included patients who were attending a family medicine clinic, our findings were likely widely relevant and applicable in general clinical practice. With heart failure and, generally, cardiovascular diseases becoming more prevalent,(4) monitoring of cholesterol variability may be an attractive and viable option for risk stratification in primary care, facilitating and optimizing primary prevention of these conditions. Third, and extending from the above, our findings may encourage further research into interventions, whether pharmacological or behavioural, that optimize cholesterol variability. Such evidence appears to be extremely scarce at the moment. Given our findings, further research in this area is strongly warranted.

### Limitations

This study has several limitations. First, only 4% of all patients attending a family clinic (5662 of 155,065) were included in this study, with the majority (92.9%) having been excluded due to having less than three sets of fasting lipid profile test results. As such, there may be selection bias for those who had indications for multiple lipid profile testing, which may translate to a higher baseline risk than the general population. This may limit the generalizability of our results. Second, since echocardiographic data is not available from CDARS, we had no information about the aetiology and subtype of heart failure that were diagnosed during the study period. As the pathophysiology of heart failure varies considerably with different aetiologies and subtypes, further studies that capture these information would be useful in further elucidating the association of cholesterol variability with the risks of different types of heart failure. Third, CDARS only recorded prescription data, but not medication adherence. The effects of medication non-adherence on lipid variability and the prognostic values of these measures of variability could not be estimated. Nonetheless, our subgroup analyses showed substantial differences between subgroups, suggesting that usage of lipid-lowering medication(s) indeed had significant effects on the variability measures’ prognostic values. Fourth, since all diagnoses were identified using ICD-9 codes as recorded by CDARS, the data could not be adjudicated individually. Nonetheless, the data input was performed by the treating physicians, and none of the authors had influence over these inputs. Data from CDARS has also been used extensively in other studies.(16,26,39) Fifth and lastly, the observational nature of this study implied that there may be other significant confounders that we have not accounted for, which may include other comorbidities, smoking status, body composition, or intercurrent illnesses. Nonetheless, we have adjusted for a range of well-established cardiovascular risk factors in the multivariable Cox regression analyses, which should account for most potential confounding factors pertinent to our outcomes.

## Conclusion

Higher HDL-C variability was associated with higher risks of adverse heart failure. Higher LDL-C and HDL-C variability were associated with higher risk of cardiovascular mortality, but not risk of myocardial infarction. These associations may be negated by usage of lipid-lowering medication(s). Further studies into the relationships between cholesterol variability, inflammation and heart failure, as well as interventions reducing cholesterol variability are warranted.

## Data Availability

All data produced in the present study are available upon reasonable request to the authors

