## Supplementary Table for "High visit-to-visit cholesterol variability predicts heart failure and adverse cardiovascular events: a population-based cohort study"

**Supplementary Material**

**Supplementary Table 1.** ICD-9 codes used to identify outcomes and co-morbidities.

| Heart failure | 428 428 428.1 428.2 428.2 428.21 428.22 428.23 428.3 428.3 428.31 428.32 428.33 428.4 428.4 428.41 428.42 428.43 428.9 398.91 402.01 402.11 402.91 404.01 404.03 404.11 404.13 404.91 404.93 |
| --- | --- |
| Myocardial infarction | 410 |
| Diabetes mellitus | 250 250.01 250.02 250.03 250.1 250.11 250.12 250.13 250.2 250.21 250.22 250.23 250.3 250.31 250.32 250.33 250.4 250.41 250.42 250.43 250.5 250.51 250.52 250.53 250.6 250.61 250.62 250.63 250.7 250.71 250.72 250.73 250.8 250.81 250.82 250.83 250.9 250.91 250.92 250.93 |
| Hypertension | 401 401.1 401.9 402 402.01 402.1 402.11 402.9 402.91 403 403.01 403.1 403.11 403.9 403.91 404 404.01 404.02 404.03 404.1 404.11 404.12 404.13 404.9 404.91 404.92 404.93 405 405.01 405.09 405.1 405.11 405.19 405.9 405.91 405.99 437.2 |
| Atrial fibrillation | 427.31 429.4 |
| Stroke / transient ischaemic attack | 435 435.1 435.2 435.3 435.8 435.9 433.81 433.91 434 436 437 437.1 433.31 433.01 434.01 434.1 434.11 434.9 434.91 437.2 437.3 437.4 437.5 437.6 437.7 437.8 437.9 430 431 432 432.1 432.9 |
| Dementia or Alzheimer’s disease | 331.82 290 290.1 290.11 290.12 290.13 290.2 290.21 290.3 290.4 290.41 290.42 290.43 290.8 290.9 294.2 294.1 294.11 294.21 332 46.1 333.4 340 42331 331.19 294.29 |
| Chronic obstructive pulmonary disease | 490 491 492 493 494 495 496 491.1 491.2 491.21 491.22 491.8 491.9 492.8 493.01 493.02 493.1 493.11 493.12 493.2 493.21 493.22 493.8 493.81 493.82 493.9 493.91 493.92 494.1 495.1 495.2 495.3 495.4 495.5 495.6 495.7 495.8 495.9 |
| Peripheral vascular disease | 250.7 443.9 443 443.1 443.2 443.21 443.22 443.23 443.24 443.29 443.8 443.81 443.82 443.89 441 443.9 785.4 V43.4 |
| Ischaemic heart disease | 410.01 410.02 410.1 410.11 410.12 410.2 410.21 410.22 410.3 410.31 410.32 410.4 410.41 410.42 410.5 410.51 410.52 410.6 410.61 410.62 410.7 410.71 410.72 410.8 410.81 410.82 410.9 410.91 410.92 411 411.1 411.8 411.81 411.89 413 413.1 413.9 414 414.01 414.02 414.03 414.04 414.05 414.06 414.07 414.1 414.11 414.12 414.19 414.2 414.3 414.4 414.8 414.9 410 412 |

**Supplementary Table 2.** Results of univariable Cox regression analysis for the overall study cohort. Numbers shown are hazard ratios with 95% confidence intervals.

|  | New-onset heart failure | Cardiovascular mortality | Myocardial infarction |
| --- | --- | --- | --- |
| Age | 1.093 [1.087, 1.100], p<0.0001 | 1.103 [1.092, 1.113], p<0.0001 | 1.077 [1.068, 1.086], p<0.001 |
| Male sex | 1.054 [0.937, 1.185], p=0.383 | 1.317 [1.111, 1.561], p=0.001 | 1.447 [1.240, 1.688], p<0.0001 |
| Hypertension | 1.709 [1.478, 1.976], p<0.0001 | 1.618 [1.31, 2.013], p<0.0001 | 1.502 [1.225, 1.842], p<0.0001 |
| Diabetes mellitus | 1.677 [1.492, 1.885], p<0.0001 | 1.573 [1.322, 1.872], p<0.0001 | 1.897 [1.623, 2.218], p<0.0001 |
| Atrial fibrillation | 2.588 [1.664, 4.028], p<0.0001 | 2.561 [1.370, 4.788], p=0.003 | 0.418 [0.104, 1.676], p=0.218 |
| Chronic obstructive pulmonary disease | 2.514 [1.423, 4.441], p=0.001 | 1.877 [0.701, 5.022], p=0.210 | 2.385 [1.067, 5.331], p=0.034 |
| Ischaemic heart disease | 1.952 [1.572, 2.423], p<0.0001 | 1.548 [1.094, 2.191], p=0.014 | 2.077 [1.568, 2.752], p<0.0001 |
| Peripheral vascular disease | 3.263 [1.627, 6.541], p=0.001 | 5.399 [2.414, 12.075], p<0.0001 | 6.140 [2.913, 12.943], p<0.0001 |
| Stroke or transient ischaemic attack | 1.833 [1.276, 2.634], p=0.001 | 2.603 [1.647, 4.115], p<0.0001 | 2.050 [1.283, 3.276], p=0.003 |
| Dementia or Alzheimer’s disease | 2.276 [0.853, 6.075], p=0.101 | 4.054 [1.302, 12.619], p=0.016 | 1.086 [0.153, 7.727], p=0.934 |
| Mean LDL | 0.961 [0.878, 1.053], p=0.393 | 1.327 [1.169, 1.506], p<0.0001 | 0.874 [0.771, 0.991], p=0.035 |
| SD of LDL | 1.050 [0.869, 1.270], p=0.612 | 0.865 [0.649, 1.153], p=0.323 | 1.360 [1.060, 1.745], p=0.016 |
| CV of LDL | 0.748 [0.431, 1.300], p=0.304 | 0.101 [0.043, 0.237], p<0.0001 | 2.887 [1.401, 5.951], p=0.004 |
| Mean HDL | 0.546 [0.456, 0.654], p<0.0001 | 0.482 [0.367, 0.633], p<0.0001 | 0.394 [0.305, 0.508], p<0.0001 |
| SD of HDL | 5.190 [3.162, 8.518], p<0.0001 | 2.827 [1.251, 6.388], p=0.012 | 5.644 [2.873, 11.086], p<0.0001 |
| CV of HDL | 70.74 [31.583, 158.446], p<0.0001 | 9.713 [2.529, 37.305], p=0.001 | 182.114 [64.480, 514.352], p<0.0001 |

CV, coefficient of variation. HDL-C, high density lipoprotein cholesterol. LDL-C, low density lipoprotein cholesterol. SD, standard deviation.

**Supplementary Table 3.** Results of univariable Cox regression analysis for patients without baseline usage of lipid-lowering drug(s). Numbers shown are hazard ratios with 95% confidence intervals.

|  | New-onset heart failure | Cardiovascular mortality | Myocardial infarction |
| --- | --- | --- | --- |
| Age | 1.095 [1.087, 1.103], p<0.0001 | 1.100 [1.088, 1.113], p<0.0001 | 1.078 [1.068, 1.089], p<0.0001 |
| Male sex | 1.056 [0.915, 1.218], p=0.456 | 1.402 [1.133, 1.735], p=0.002 | 1.425 [1.178, 1.725], p=0.0003 |
| Hypertension | 1.889 [1.584, 2.252], p<0.0001 | 1.804 [1.374, 2.368], p<0.0001 | 1.588 [1.233, 2.047], p=0.0003 |
| Diabetes mellitus | 1.681 [1.455, 1.943], p<0.0001 | 1.680 [1.347, 2.096], p<0.0001 | 1.661 [1.362, 2.026], p<0.0001 |
| Atrial fibrillation | 2.256 [1.276, 3.989], p=0.005 | 3.822 [1.971, 7.411], p<0.0001 | 0.331 [0.047, 2.356], p=0.270 |
| Chronic obstructive pulmonary disease | 2.984 [1.645, 5.414], p=0.0003 | 1.932 [0.619, 6.025], p=0.257 | 2.640 [1.092, 6.381], p=0.031 |
| Ischaemic heart disease | 2.108 [1.528, 2.908], p<0.0001 | 2.302 [1.449, 3.656], p=0.0004 | 2.486 [1.660, 3.725], p<0.0001 |
| Peripheral vascular disease | 3.045 [1.140, 8.134], p=0.026 | 5.066 [1.625, 15.790], p=0.005 | 6.510 [2.430, 17.438], p=0.0002 |
| Stroke or transient ischaemic attack | 2.288 [1.468, 3.567], p=0.0003 | 3.533 [2.029, 6.151], p<0.0001 | 2.382 [1.309, 4.336], p=0.004 |
| Dementia or Alzheimer’s disease | 3.688 [1.187, 11.463], p=0.024 | 8.807 [2.823, 27.479], p=0.0002 | 2.242 [0.315, 15.961], p=0.420 |
| Mean LDL | 0.931 [0.831, 1.042], p=0.931 | 1.207 [1.020, 1.428], p=0.029 | 0.883 [0.755, 1.033], p=0.120 |
| SD of LDL | 0.953 [0.749, 1.213], p=0.699 | 0.763 [0.522, 1.115], p=0.162 | 1.351 [0.980, 1.863], p=0.066 |
| CV of LDL | 0.587 [0.300, 1.149], p=0.120 | 0.100 [0.034, 0.291], p<0.0001 | 2.558 [1.041, 6.283], p=0.040 |
| Mean HDL | 0.581 [0.468, 0.721], p<0.0001 | 0.414 [0.292, 0.588], p<0.0001 | 0.443 [0.325, 0.604], p<0.0001 |
| SD of HDL | 5.776 [3.229, 10.331], p<0.0001 | 5.606 [2.221, 14.152], p=0.0003 | 8.945 [4.172, 19.181], p<0.0001 |
| CV of HDL | 54.945 [21.185, 142.501], p<0.0001 | 29.384 [6.274, 137.614], p<0.0001 | 218.343 [65.376, 729.229], p<0.0001 |

CV, coefficient of variation. HDL-C, high density lipoprotein cholesterol. LDL-C, low density lipoprotein cholesterol. SD, standard deviation.

**Supplementary Table 4.** Results of univariable Cox regression analysis for patients with baseline usage of lipid-lowering drug(s). Numbers shown are hazard ratios with 95% confidence intervals.

|  | New-onset heart failure | Cardiovascular mortality | Myocardial infarction |
| --- | --- | --- | --- |
| Age | 1.089 [1.076, 1.103], p<0.0001 | 1.107 [1.088, 1.127], p<0.0001 | 1.072 [1.055, 1.088], p<0.0001 |
| Male sex | 1.046 [0.852, 1.286], p=0.666 | 1.187 [0.896, 1.574], p=0.232 | 1.489 [1.144, 1.937], p=0.003 |
| Hypertension | 1.341 [1.036, 1.735], p=0.026 | 1.262 [0.876, 1.818], p=0.212 | 1.289 [0.915, 1.815], p=0.147 |
| Diabetes mellitus | 1.566 [1.281, 1.914], p<0.0001 | 1.281 [0.966, 1.699], p=0.086 | 2.226 [1.713, 2.892], p<0.0001 |
| Atrial fibrillation | 3.407 [1.690, 6.868], p=0.001 | 0.626 [0.088, 4.470], p=0.641 | 0.549 [0.077, 3.913], p=0.549 |
| Chronic obstructive pulmonary disease | 1.100 [0.155, 7.831], p=0.924 | 2.235 [0.313, 15.949], p=0.423 | 1.985 [0.278, 14.155], p=0.494 |
| Ischaemic heart disease | 1.612 [1.197, 2.171], p=0.002 | 0.853 [0.504, 1.444], p=0.554 | 1.501 [1.010, 2.232], p=0.044 |
| Peripheral vascular disease | 3.183 [1.188, 8.531], p=0.021 | 5.088 [1.625, 15.934], p=0.005 | 5.220 [1.667, 16.347], p=0.005 |
| Stroke or transient ischaemic attack | 1.188 [0.634, 2.227], p=0.590 | 1.426 [0.633, 3.213], p=0.392 | 1.491 [0.703, 3.166], p=0.298 |
| Dementia or Alzheimer’s disease | 0.947 [0.133, 6.742], p=0.957 | 0.050 [0.000, 17631.549], p=0.645 | 0.050 [0.000, 7683.465], p=0.622 |
| Mean LDL | 0.972 [0.834, 1.132], p=0.713 | 1.403 [1.160, 1.697], p=0.0005 | 0.807 [0.654, 0.995], p=0.044 |
| SD of LDL | 1.011 [0.733, 1.394], p=0.947 | 0.733 [0.464, 1.159], p=0.184 | 1.071 [0.706, 1.626], p=0.747 |
| CV of LDL | 0.770 [0.281, 2.108], p=0.610 | 0.036 [0.008, 0.160], p<0.0001 | 2.052 [0.579, 7.280], p=0.266 |
| Mean HDL | 0.496 [0.358, 0.687], p<0.0001 | 0.655 [0.424, 1.010], p=0.055 | 0.325 [0.207, 0.510], p<0.0001 |
| SD of HDL | 3.982 [1.528, 10.377], p=0.005 | 0.474 [0.088, 2.558], p=0.385 | 1.554 [0.374, 6.462], p=0.544 |
| CV of HDL | 144.291 [29.165, 713.873], p<0.0001 | 0.349 [0.022, 5.418], p=0.452 | 107.873 [12.842, 906.117], p<0.0001 |

CV, coefficient of variation. HDL-C, high density lipoprotein cholesterol. LDL-C, low density lipoprotein cholesterol. SD, standard deviation.
